# A large expert-annotated single-cell peripheral blood dataset for hematological disease diagnostics

**DOI:** 10.1101/2025.02.18.25322415

**Authors:** Sayedali Shetab Boushehri, Armin Gruber, Salome Kazeminia, Christian Matek, Karsten Spiekermann, Christian Pohlkamp, Torsten Haferlach, Carsten Marr

**Affiliations:** Institute of AI for Health, Helmholtz Munich – German Research Center for Environmental Health, Neuherberg, Germany; Data & Analytics, Pharmaceutical Research and Early Development (pRED), Roche Innovation Center Munich (RICM), Penzberg, Germany; TUM School of Computation, Information and Technology, Technical University of Munich, Munich, German; Laboratory of Leukemia Diagnostics, Department of Medicine III, University Hospital, LMU Munich, Munich, Germany; German Cancer Consortium (DKTK), Heidelberg, Germany; German Cancer Research Center (DKFZ), Heidelberg, Germany; Munich Leukemia Laboratory, Munich, Germany

**Keywords:** single-cell images, white blood cell, AI, hematology, cytomorphology, leukemia

## Abstract

Distinguishing cell types in peripheral blood smears is critical for diagnosing blood diseases, such as leukemia subtypes. Artificial intelligence can assist in automating cell classification. For training robust machine learning algorithms, however, large and well-annotated single-cell datasets are pivotal.

Here, we introduce a large, publicly available, annotated peripheral blood dataset comprising >40,000 single-cell images classified into 18 classes by cytomorphology experts from the Munich Leukemia Laboratory, the largest European laboratory for blood disease diagnostics. By making our dataset publicly available, we provide a valuable resource for medical and machine learning researchers and support the development of reliable and clinically relevant diagnostic tools for diagnosing hematological diseases.

## Background & Summary

Microscopic examination and classification of blood cells play a crucial role in diagnosing hematological diseases. This process involves evaluating the morphology of leukocytes and is typically performed by human experts trained over the years. Like other diagnostic tasks, it is repetitive, time-consuming, and susceptible to intra- and inter-observer variation^1^. One promising solution is the development of automatic single-cell classifiers using machine learning, which can substantially reduce the time and effort required by experts^2^. Deep learning, in particular, has been used for diagnosing hematological diseases from single-cell images in peripheral blood^3–9^ and bone marrow^10–12^.

As supervised deep learning crucially relies on large amounts of annotated data, a current lack of large datasets creates a bottleneck for improving the accuracy of classifiers^13^. This work presents the largest publicly available, expert-annotated dataset of peripheral blood single-cells, with over 40,000 images. While our dataset is being published here for the first time, it has been used in previous studies^4,5,19–22^.

## Methods

The data acquisition process by the Munich Leukemia Laboratory took several steps (see also Hehr et al.^4^). Blood samples and smears were collected between 2000 and 2020 from patients with a wide distribution of hematological diagnoses. A patient cohort with blood samples from adult patients who gave informed consent to the use of their data for research purposes was selected. Blood smears were scanned with a 10x objective for an overview image. Cell detection was performed using the Metasystems Metafer software. After applying a segmentation threshold and a logarithmic color transformation, stained cells with an object size between 40–800 μm^2^ were detected and stored in a gallery. Each gallery image was assigned to a quality level using a classifier to determine the cell density and immediate cell neighborhood. Cell imaging and detection were then performed in 40x on high-quality cells using a segmentation threshold, a logarithmic color transformation, and an object size between 40–800 μm^2^. The resulting 41,906 images of single nucleated cells comprise 288 × 288 pixels and 25μm x 25μm, corresponding to a resolution of 11.52 pixels per μm. Subsequently, five human expert examiners at the Munich Leukemia Laboratory annotated the images, assigning each single cell to one out of 18 classes (Fig. 1a).

**Figure 1:**
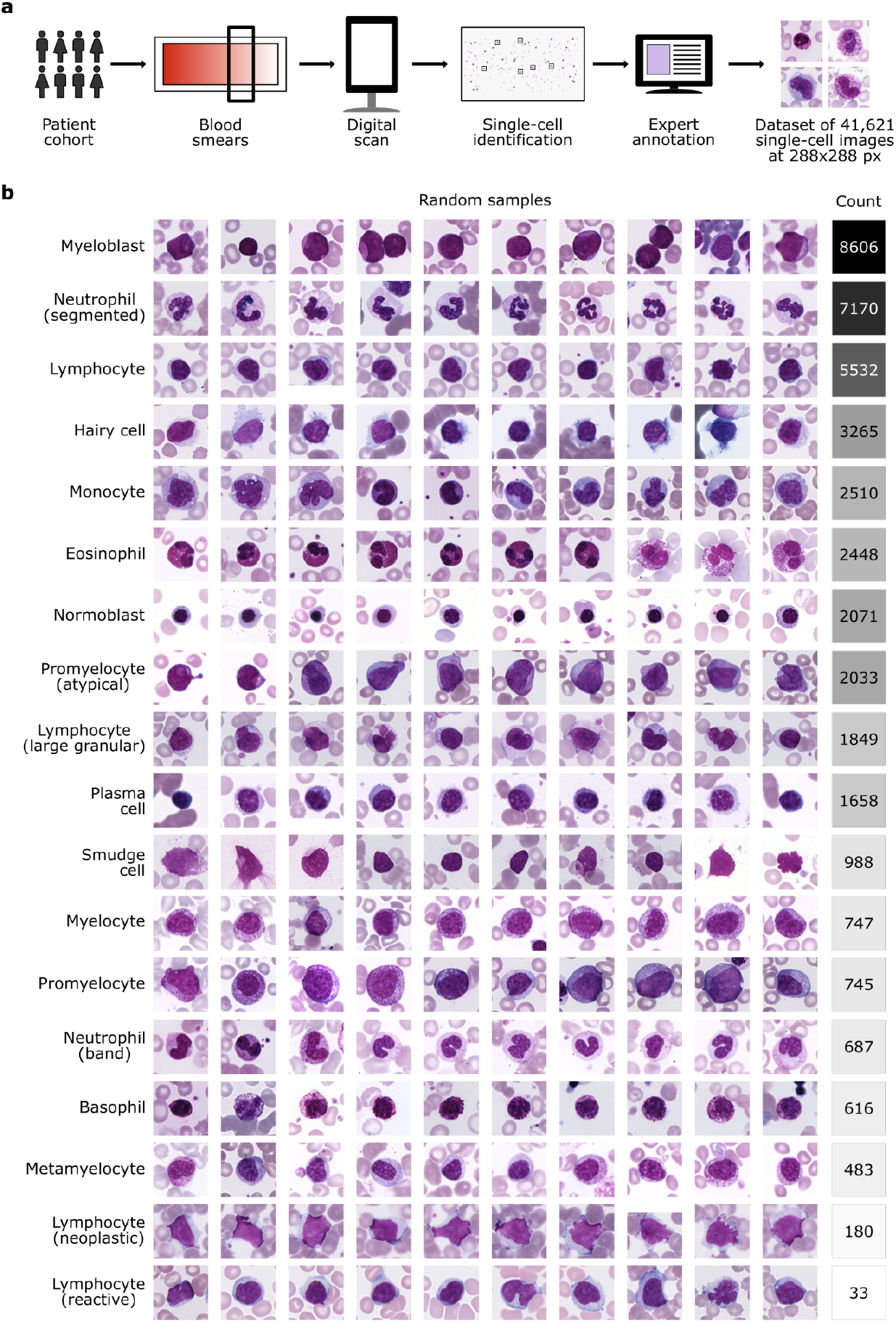
A fully annotated single-cell peripheral blood dataset. **a** Workflow of generating the imaging dataset at the Munich Leukemia Laboratory **b** The MLL23 dataset contains 18 classes with varying numbers of images per class. Ten representative images per class are depicted to provide an overview of the dataset.

We reduced the dataset to 41,621 cells by deleting duplicate images. Some duplicate images also had differing labels, corresponding to indecisive borderline cases. Note that some cells are depicted in two or more images, but with differing focus or cropping.

In the group of lymphoid cells, there are mature ‘typical lymphocytes’ (number of single-cell images = 5,532) and atypical lymphocytes like plasma cells (1,658), large granular lymphocytes (1,849), reactive lymphocytes (33), hairy cells (3,265) and other neoplastic lymphocytes (180), as well as smudge cells (988). In comparison, the group of myeloid cells is divided into mature cells like band neutrophil granulocytes (687), segmented neutrophil granulocytes (7,170), eosinophil granulocytes (2,448), basophil granulocytes (616), monocytes (2510), and immature cells like myeloblasts (8,606), metamyelocytes (483), promyelocytes (745), myelocytes (747), and atypical promyelocytes (2,033). Lastly, normoblasts (2071) are also present in the dataset. The cell types occur with specific frequencies in the peripheral blood in healthy and pathological patients. Due to the Munich Leukemia Laboratory’s focus on hematologic neoplasms, the dataset is inherently imbalanced in terms of the number of images per class. For instance, it contains over 8,000 myeloblasts but only 33 reactive lymphocytes (Fig. 1b).

## Data Records

The dataset is hosted on Zenodo and can be accessed via https://zenodo.org/uploads/14277609. It comprises 18 ZIP files, each named after a specific cell type (e.g., basophil.zip). Each ZIP file contains high-quality TIFF images of individual cells belonging to the corresponding class, with file names following a consistent format that includes the class name and a unique identifier (e.g., basophil_0001.TIF).

## Ethics declaration

Informed consent was obtained indirectly, at the time of routine collection, for possible research. All patients in the MLL23 dataset were at least 18 years old. Ethics approval was granted by the Ethics Committee of LMU Munich (reference number 19-696).

## Data Availability

Zenodo

https://zenodo.org/uploads/14277609

## Acknowledgments

The authors thank Xudong Sun, Matthias Hehr, Sophia J. Wagner, Valentin Koch, and Matteo Wohlrapp (all from Munich) for the fruitful discussions.

## Author contributions

ChM conceived the project idea with CM. SSB and AG performed the data cleaning, wrote the manuscript, and designed the figures with CM. CM supervised the study with KS. SK helped with the manuscript consistency and edits. CP and TH performed main data collection, annotation, and pseudonymization.

